# Optimal control of an SIR epidemic through finite-time non-pharmaceutical intervention

**DOI:** 10.1101/2020.05.05.20091439

**Authors:** David I. Ketcheson

**Affiliations:** King Abdullah University of Science & Technology

**Author notes:** Computer, Electrical, and Mathematical Sciences & Engineering Division, King Abdullah University of Science and Technology, 4700 KAUST, Thuwal 23955, Saudi Arabia.

## Abstract

We consider the problem of controlling an SIR-model epidemic by temporarily reducing the rate of contact within a population. The control takes the form of a multiplicative reduction in the contact rate of infectious individuals. The control is allowed to be applied only over a finite time interval, while the objective is to minimize the total number of individuals infected in the long-time limit, subject to some cost function for the control. We first consider the no-cost scenario and analytically determine the optimal control and solution. We then study solutions when a cost of intervention is included, as well as a cost associated with overwhelming the available medical resources. Examples are studied through the numerical solution of the associated Hamilton-Jacobi-Bellman equation. Finally, we provide some examples related directly to the current pandemic.

**AMS subject classification**. 92D30, 34H05, 49N90, 92-10, 49L12

## 1 Problem description and assumptions

The classical SIR model of Kermack & Mckendrick [13] is

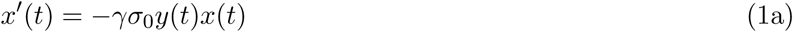

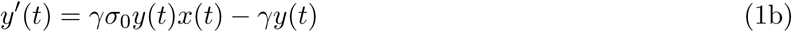

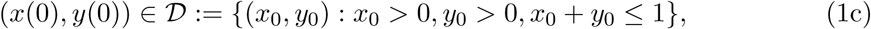

where *x*(*t*)*,y*(*t*) represent the susceptible and infected populations respectively, while the recovered population is *z*(*t*) = 1 − *x*(*t*) − *y*(*t*). The region 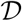 is forward-invariant and a unique solution exists for all time [10]. While the temporal dynamics of (1) depend on both *σ*_0_ and *γ*, the set of trajectories depends only on the basic reproduction number *σ*_0_. Dynamics for two values of *σ*_0_ are shown in Figure 1.

**Figure 1:**
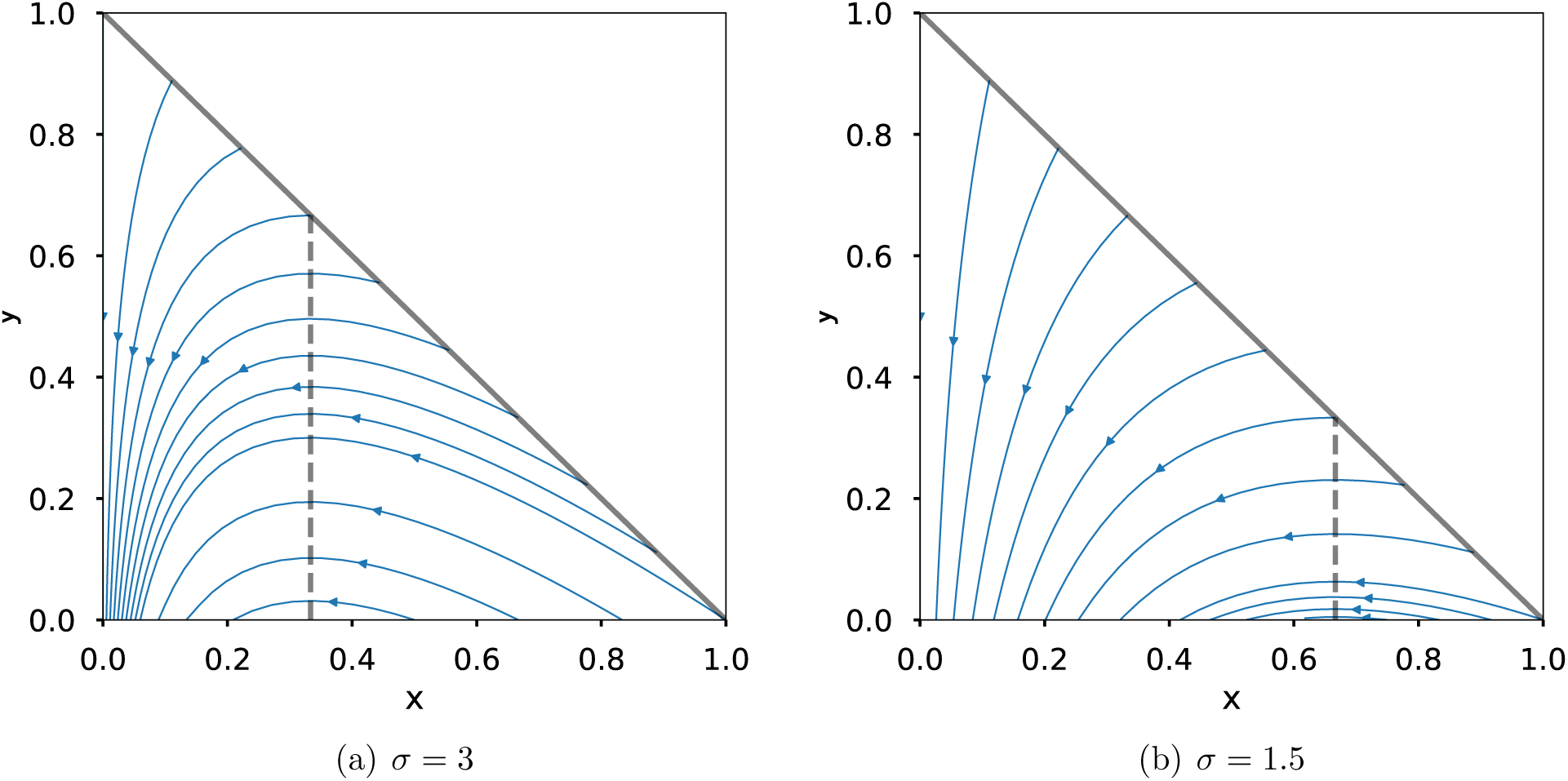
Dynamics of the SIR model (1) for two values of the basic reproduction number. The critical value *x* = 1/*σ* is shown with a dashed line.

The system (1) is at equilibrium if *y*(*t*) = 0. This equilibrium is stable if and only if *x*(*t*) ≥ 1/*σ*_0_, a condition referred to as *herd immunity*. If this condition is not satisfied at the initial time, then *y*(*t*) will first increase until it is, and then decrease, approaching zero asymptotically. The SIR model assumes that recovery confers permanent immunity.

For many diseases affecting humans, herd immunity is achieved through vaccination of a sufficient portion of the population. Herein we assume a vaccine is unavailable, so that herd immunity can only be achieved through infection and recovery. Our goal is to minimize *z*_∞_ := lim_t→∞_ *z*(*t*), or equivalently (since *y_∞_* = 0) to maximize the long-time limit of the susceptible fraction: *x_∞_* = lim_t→_*_∞_x*(*t*). This has the effect of minimizing the number of eventual deaths, which would be proportional to *z_∞_*.

This is equivalent to minimizing the number of deaths, if we assume that some fixed fraction of the recovered population *z*(*t*) dies from the disease. From the foregoing it is clear that *x_∞_* ≤ 1/*σ*_0_. The difference *1/σ*_0_ *− x_∞_* is referred to as *epidemiological overshoot*. For COVID-19, a review of early estimates of *σ*_0_ can be found in [16, Table 1], with mean 3.28 and median 2.79. In accordance with these estimates, we use a value *σ*_0_ = 3 in most of the examples in this work. With this value, the SIR model implies that eventually at least two-thirds of the world population will eventually have COVID-19 antibodies; this number is likely to be significantly higher in reality due to epidemiological overshoot. For instance, it can be seen from Figure 1(a) that, starting from a fully susceptible population and a small number of infected individuals, in the absence of control the SIR model predicts that over 90% of the population would be infected.

This overshoot can be reduced through non-pharmaceutical intervention (NPI), which is simply a means to reduce contact between infected and susceptible individuals; reductions of this kind occurred for instance as a result of NPIs imposed during the 1918 flu pandemic [3]. We model a NPI control via a time-dependent reproduction number *σ*(*t*) *∈* [0, *σ*_0_] with the system

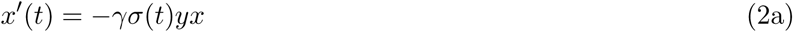

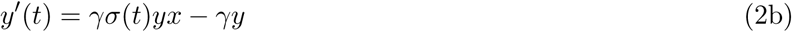

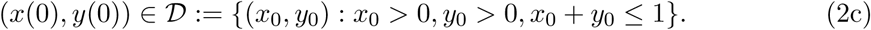

A temporary reduction in *σ* can account for both population-wide interventions and interventions specific to identified infectious (or possibly infectious) individuals. The SIR model with a time-dependent reproduction number (or equivalently, a time-dependent contact rate) has been considered before, for instance in [3, 22].

Typically, an epidemic does not result in substantial *permanent* change in the contact rate of a population. We therefore assume

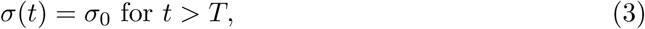

i.e., that intervention can only be applied over a finite interval *t* ∈ [0, *T*]. Since *x*_∞_ = 1/*σ*_0_ only at the single point (*x* = 1/*σ*_0_,*y* = 0), and since the *y* = 0 axis cannot be reached in a finite time, (3) implies that any solution must have *x*_∞_ < 1/*σ*_0_.

We state the control problem as follows:

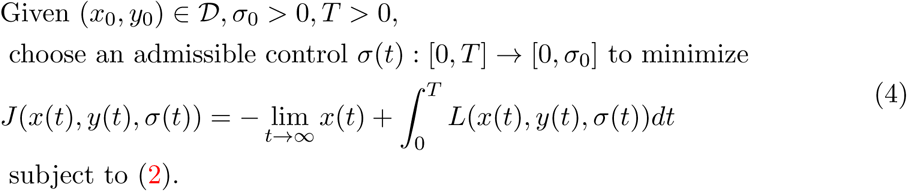

Here *J* is the objective function that accounts for the desire to minimize infections as well as a running cost of imposing control. We assume throughout that *L* is convex with respect to *q*(*t*) = 1 − *σ*/*σ*_0_.

There is a large body of work on compartmental epidemiological models and control for such models; see e.g. [10, 15] and references therein. A number of works focus on optimal control through vaccination; see e.g. [12]. Other works, such as [25, 20, 1] focus on explicit modeling of and/or control through quarantined and isolated individuals. A review of work on optimal control in compartmental epidemiological models is presented in [21], along with the formulation of necessary conditions (based on Pontryagin’s maximum principle) for various extensions of the SIR model. For modeling and control based on even more detailed models incorporating spatial spread and human networks, see e.g. [4].

### 1.1 Objectives and contributions

The modeling and assumptions in the present work are motivated by the current COVID-19 epidemic, which so far is being managed through broad NPIs and without a vaccine. In order to understand the effects of NPIs imposed on an entire population, we stick to the simple model (2) rather than explicitly modeling quarantined individuals. Since such population-wide measures cannot be maintained indefinitely, we invoke the finite-time control assumption (3). This assumption is not new (see e.g. [7]), but unlike previous works our objective function is still based on the long-term outcome (rather than the outcome at time *T*). This drastically changes the nature of optimal solutions.

Although the broad motivation for this work comes from the current epidemic, our primary objective is to understand general properties of optimal controls for the variable-*σ* SIR system (2). To this end, we also investigate solutions in certain asymptotic regimes (such as when there is little or no cost associated with the control). Nevertheless, the values of the key parameters *γ* and *σ*_0_ for all examples are chosen to fall in the range of current estimates for COVID-19.

One novel aspect of this work is that the problem is posed in terms of the infinite-time limit, but formulated in a way that only requires solution over a finite time interval. Indeed, without this reformulation we found that the problem was extremely ill-conditioned; this reformulation is also needed in order to compute approximate solutions via a Hamilton-Jacobi-Belmman equation. This reformulation is presented in Section 2. The main theoretical result is an exact characterization of the optimal control when *L* = 0, given as Theorem 3 in Section 3.

Typical results in the literature on control of compartmental epidemiological models are numerical and are based on Pontryagin’s weak maximum principle, which gives only necessary conditions for optimality. At best, uniqueness is shown for small times; see e.g. [14, 5, 11, 25, 12, 21]. In contrast, here the main result includes a proof of optimality for arbitrarily large times. In Section 4 we explore the behavior of optimal solutions for *L* ≠ 0 under various interesting cost functions and parameter regimes. Here the results are based on solutions of the relevant Hamilton-Jacobi-Bellman equation, which is both necessary and sufficient for optimality. In Section 5 we consider direct application to the COVID-19 pandemic. Some conclusions are drawn in Section 6.

## 2 Formulation over a finite time interval

In this section we reformulate the control problem (4) in terms of the solution over a finite time interval [0, *T*]. This reformulation is necessary both to facilitate the exact solution in Section 3 and to arrive at a numerically-tractable problem for computing approximate solutions, as described in Section 4.

In general, the solution of (2) depends on the initial data (*x*_0_, *y*_0_), the control *σ*(*t*), and time *t*, so it is natural to write *x*(*t*; *σ*(*t*)*,x*_0_, *y*_0_). In what follows it will be convenient to make a slight abuse of notation and write *x*(*t*; *σ*(*t*)) or *x*(*t*) when there is no chance of confusion.

For a fixed reproduction number, the asymptotic susceptible fraction *x_∞_* can be obtained from the solution *x*(*t*)*, y*(*t*) at any time *t*, since solutions of (1) move along contours of *x*_∞_. Thus we will write *x_∞_*(*x*, *y*) or *x_∞_*(*x,y,σ*_0_).

### 2.1 A formula for *x*_∞_

In this subsection we review the solution of the SIR model without control (1). It can be shown that *x*(*t*) satisfies (see [8, 18] and [13, pp.707-708])

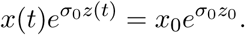

Since *z* = 1 − *x* − *y* we define

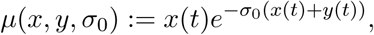

which is constant in time for any solution of (1). The trajectories in Figure 1 are thus also contours of *μ*. Since *y*_∞_ = 0, we have

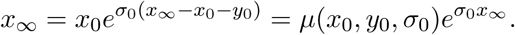

Setting *w* = −*x*_∞_*σ*_0_ we have

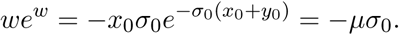

Thus *w* = *W*_0_(−*μσ*_0_) where *W*_0_ is the principal branch of Lambert’s *W*-function [18], and

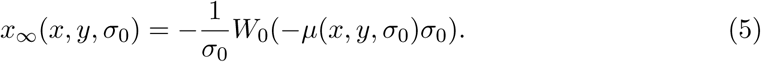

Formula (5) allows us to rewrite the problem (4) in terms of the state at time *T* < ∞: Given 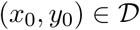, *σ*_0_ > 0, *T* > 0, choose an admissible control *σ*(*t*) : [0, *T*] → [0, *σ*_0_] to minimize

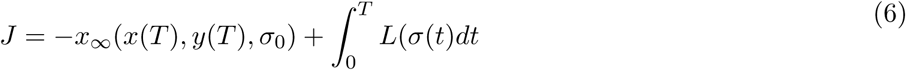

subject to (2).

In what follows we will also require the derivatives of *x*_∞_ with respect to *x*, *y*, and *μ* Direct computation gives

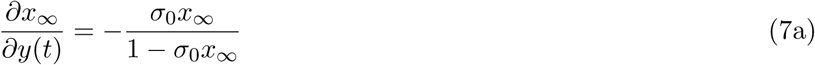

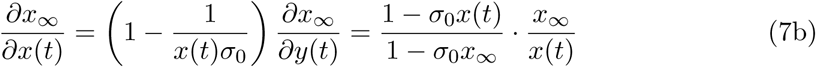

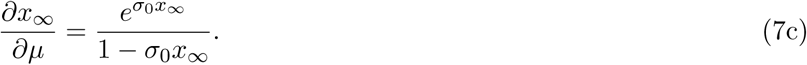

Using these expressions we can also compute the rate of change of *x*_∞_ when some control *σ*(*t*) is applied:

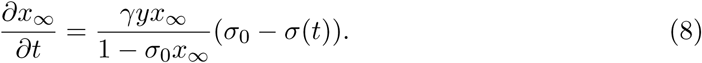

From this we see that the impact of an intervention on *x*_∞_ is independent of *x*(*t*) and directly proportional to *y*(*t*). This indicates that intervention is more impactful when there is a larger infected population.

### 2.2 Bounds on *x*_∞_

Now we turn our attention to the SIR system with control (2).

#### Definition 1. (Admissible control)

*Given a basic reproduction number σ*_0_*, we say a control fuction σ*(*t*) *is admissible if it is Lebesgue measurable and* 0 ≤ *σ*(*t*) ≤ *σ*_0_ *for all t*.

It is straightforward to show that (2) has a unique solution for all time for any initial data in 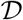 and any admissible control, by the same arguments used for (1). The proof of the next lemma shows that applying any control *σ*(*t*) *< σ*_0_ over any length of time leads to an increase in *x*_∞_.

#### Lemma 1.

*Let σ_0_ >* 0 *and* 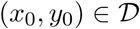 *be given. Let σ*(*t*) *be an admissible control. Then for t ≥* 0 *we have*

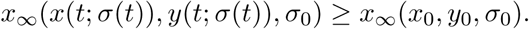

#### Proof.

Dividing (1b) by (1a) gives

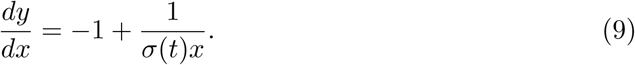

Thus reducing *σ*(*t*) has the effect of increasing *dy/dx*. Since all trajectories flow to the left (*x* is a decreasing function of *t*), this means that the solution trajectory obtained with *σ*(*t*) lies below that obtained with *σ*_0_, for all *t* > 0. Since *x_∞_* is a decreasing function of *y*, this completes the proof. □

Thus for any admissible control and any initial data we have

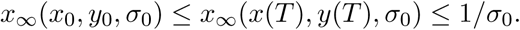

### 2.3 Existence and necessary conditions for an optimal control

Standard application of Pontryagin’s maximum principle gives necessary conditions for a solution of (6). The Hamiltonian for this problem is

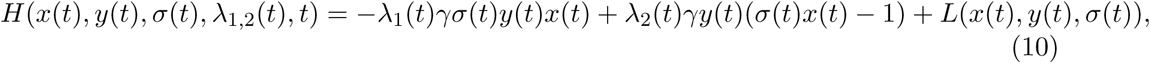

and the adjoint variables are defined by

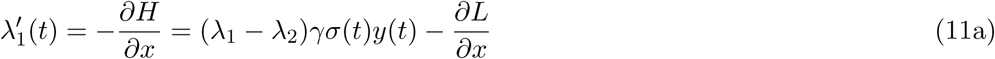

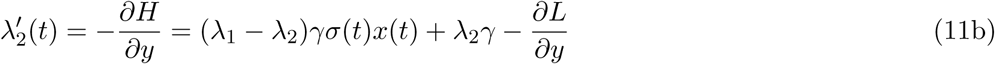

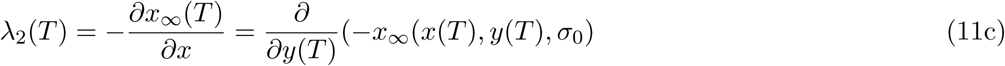

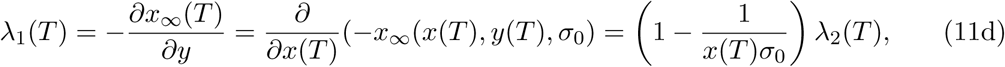

where *x*(*t*), *y*(*t*) satisfy (2). The final conditions for λ_1_,_2_ can be computed from (7).

#### Theorem 1.

*Let* 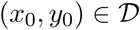 *andσ*_0_, *γ*, *T* ≥ 0 *be given. Then there exists a controlσ*^*^(*t*) *for* (6) *and corresponding response* (*x*^*^(*t*), *y*^*^(*t*)) *such thatJ is minimized over the set of admissible controls. Furthermore, there exist adjoint functionsλ*_1_,_2_(*t*) *satisfying* (11) *with x*(*t*) = *x*^*^(*t*), *y*(*t*) = *y*^*^(*t*), and the *controlσ*^*^(*t*) *satisfies the optimality condition*

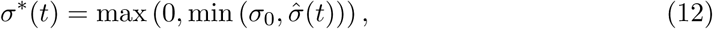

*where*

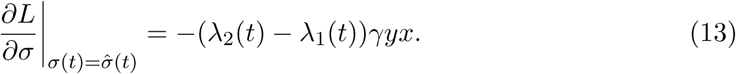

#### Proof.

The existence of an optimal control is guaranteed by [6, Corollary 4.1] since *L* is convex with respect to *q*(*t*) = 1 − *σ*(*t*)/*σ*_0_, the state solutions (*x*(*t*), *y*(*t*)) are bounded, and the system (2) is Lipschitz with respect to *x*, *y*. Thus, applying Pontryagin’s maximum principle, we convert the problem (6) into that of minimizing *H* in (10) pointwise with respect to *σ*. On the interior of the set of admissible controls we have (13), which leads to (12). □

### 2.4 Infinite-time control

In this section only, we consider controls that reach the optimal value *x*_∞_ = 1/*σ*_0_. This is achieved only at (*x*, *y*) = (1/*σ*_0_, 0), a state that cannot be reached from any other state without imposing some control, and which in any case can only be reached after an infinite time. Thus we momentarily set aside the restriction (3) and consider controls extending up to an arbitrarily large time *T*. We still require that the system approach a stable equilibrium point as *t* → ∞. We assume that *x*_0_ ≥ 1/*σ*_0_, since otherwise the maximum achievable value of *x*_∞_ is *x*_0_, which would be achieved by taking simply *σ*(*t*) = 0 for all *t*. We also take *L* = 0 so that an optimal control is any control satisfying

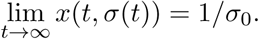

There are infinitely many such controls. Two are particularly simple and are of interest.

The first is a constant control *σ*(*t*) = *σ*_*_(*x*_0_, *y*_0_, *σ*_0_). By (5) we must have *x*_∞_(*x*_0_, *y*_0_, *σ*_*_) = 1/*σ*_0_, so *σ*_*_ is the solution of

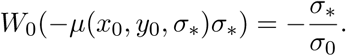

The second is a bang-bang control in which

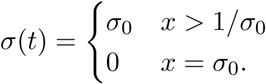

The response for each of these controls is shown for a specific example in Figure 2.

**Figure 2:**
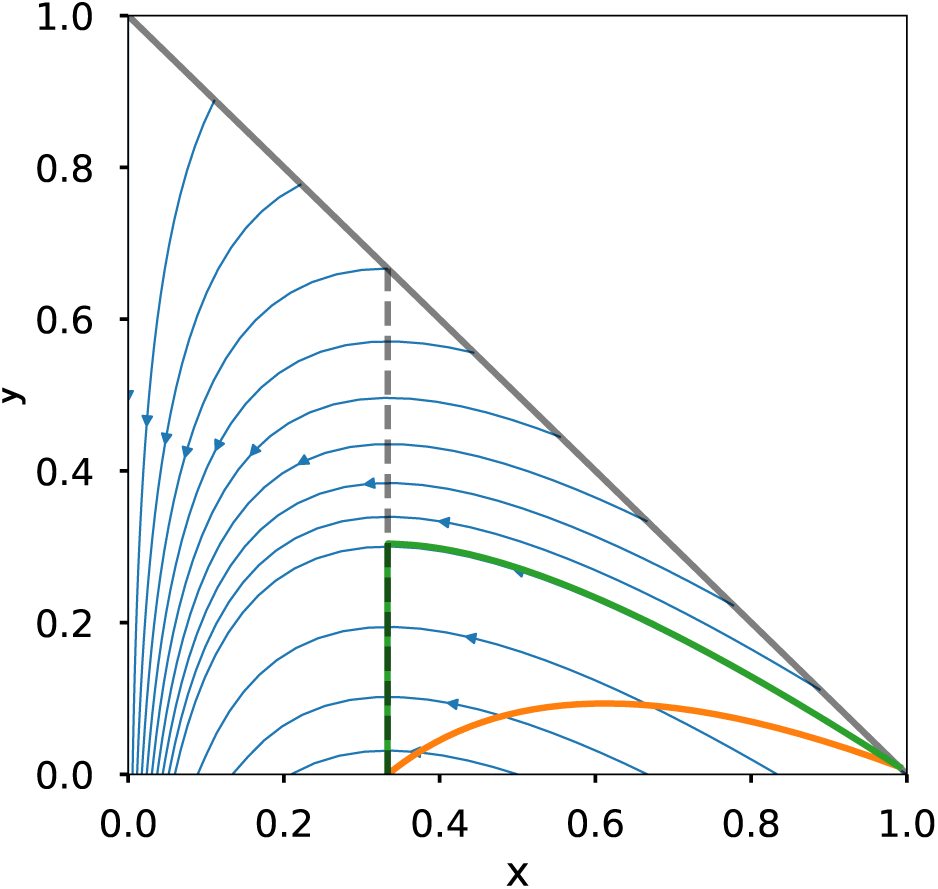
Two infinite-time controls that give *x*_∞_ = 1/*σ*_0_. Here *σ*_0_ = 3 and (*x*_0_,*y*_0_) = (0.99, 0.01). For the constant control, *σ*(*t*) = *σ_*_* ≈ (1 − 0.4557)*σ*_0_.

## 3 Optimal control with *L* = 0

In this section we derive the exact solution of the control problem (6) with *L* = 0 (i.e., when the goal of increasing *x*_∞_ completely trumps any associated costs or other concerns). Then (6) becomes

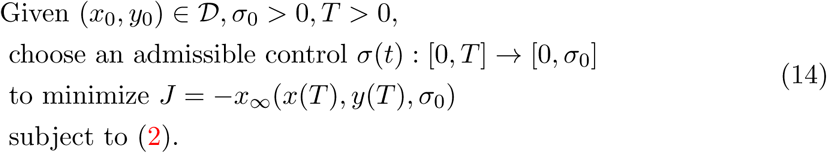

This problem can be reformulated as a minimum-time control problem.

#### Lemma 2.

*Letσ*^*^(*t*) *be an optimal control for* (14)*, and let* (*x*^*^(*T*), *y*^*^(*T*)) *denote the corresponding terminal state. Then there is no admissible control that reaches* (*x*^*^(*T*), *y*^*^(*T*)) *from* (*x*_0_,*y*_0_) *before timeT*.

#### Proof.

Suppose there were a control 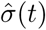 that leads to (*x*(*t*^*^), *y*(*t*^*^)) = (*x*^*^(*T*), *y*^*^(*T*)) for some *t*^*^ < *T*. Then we could obtain a smaller value of *J* in (14) by using a up to time t* combined with the choice *σ*(*t*) = 0 for *t* > *t*^*^. This contradicts the optimality of *σ*^*^(*t*). □

Furthermore, the optimal control must be a bang-bang control.

### Lemma 3.

*Letσ*(*t*) *be an optimal control for* (14)*. Then*

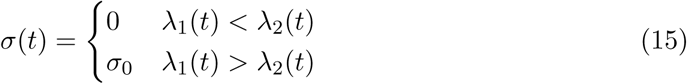

*whereλ*_1_,_2_(*t*) *are given by* (11).

### Proof.

From (10) with *L* = 0, we have

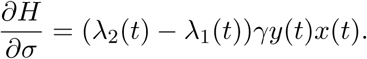

The optimality condition then implies (15) except at points where ∂*H*/∂*σ* = 0 (see e.g. [15, Ch. 17]. Since *x*(*t*), *y*(*t*) > 0 for *t* < ∞, we have that ∂H/∂*σ* = 0 if and only if *λ*_1_ = *λ*_2_. Suppose that the latter condition holds on an open interval. By (11), that would imply that 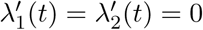 on this interval and hence for all *t*, which contradicts the boundary conditions (11). □

This motivates the following lemma.

### Lemma 4.

*Let* (*x*_0_,*y*_0_) *and* (*x*_1_,*y*_1_) *be given such thatx*_0_,*x*_1_ ≥ 1/*σ*_0_ *andx*_∞_(*x*_0_, *y*_0_, *σ*_0_) ≥ *x*_∞_(*x*_1_, *y*_1_, *σ*_0_). *Letσ*(*t*) *be a bang-bang control such that* (*x*(*t*_1_; *x*_0_, *y*_0_, *σ*(*t*)), *y*(*t*_1_; *x*_0_,*y*_0_, *σ*(*t*))) = (*x*_1_,*y*_1_) *for somet*_1_ ≥ 0. *Then the minimum value oft*_1_ *is achieved by taking*

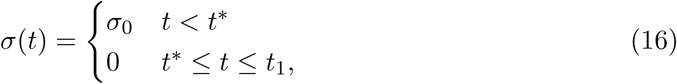

*where t^*^ satisfiesx*(*t*^*^; *x*_0_, *y*_0_, *σ*_0_) = *x*_1_.

### Proof.

Since *σ*(*t*) is a bang-bang control, the trajectory (*x*(*t*; *σ*(t)), *y*(*t*; *σ*(*t*))) consists of a sequence of segments each of which is a solution of (2) with *σ* = 0 (traveling directly downward) or with *σ* = *σ*_0_ (traveling along a contour of *x*_∞_). Some trajectories of this type are illustrated in Figure 3. Notice that each trajectory must traverse the same distance in the x-direction; since *x*′(*t*) = −*βxy* this travel is faster at larger *y* values. Meanwhile, the total length of all the downward (*σ* = 0) segments is the same for any trajectory, and since for these segments *y*′(*t*) = −*γy*, travel is again faster at larger *y* values. The control given in the lemma makes all these traversals at the largest possible values of *y*, so it arrives in the shortest time. □

Combining these three lemmas, we obtain the following.

**Figure 3:**
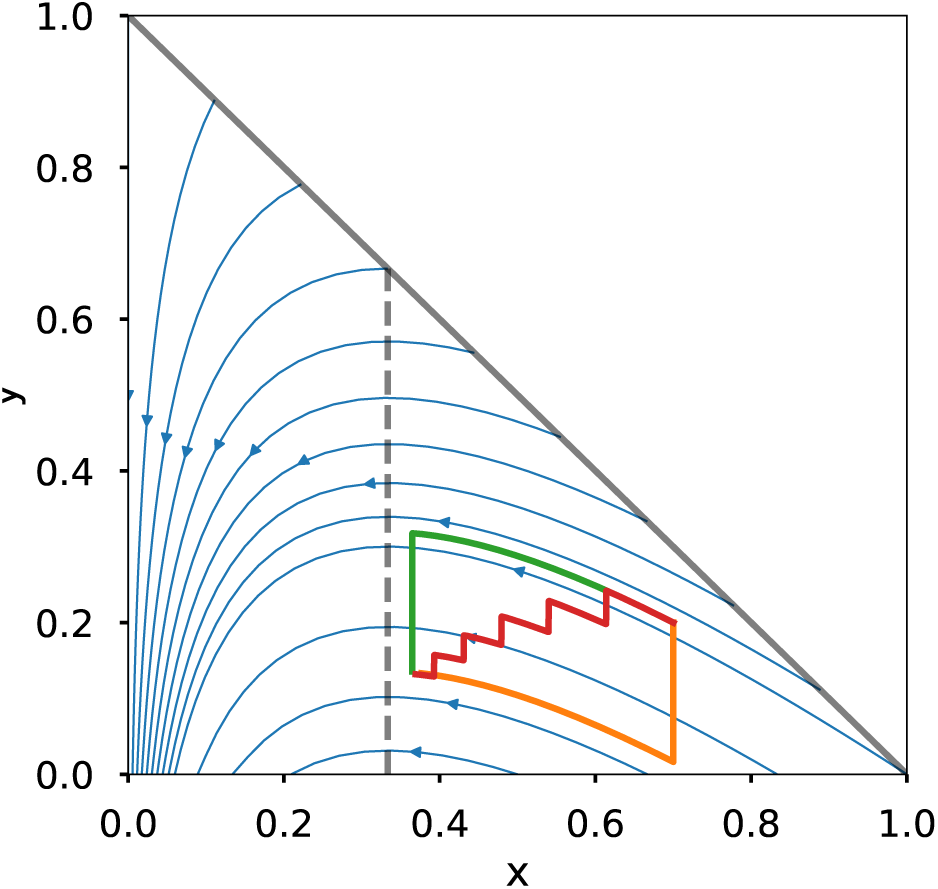
Three different paths between two states, each obtained with a bang-bang control. The top (green) path arrives in the shortest time.

### Theorem 2.

*Any optimal control for* (14) *is of the form* (16) *witht*_1_ = *T*.

### Proof.

By Lemmas 2 and 3, the optimal control must be bang-bang and must solve the optimal-time problem. Then Lemma 4 applies and gives the stated result. □

We can now give the solution of (14).

### Theorem 3.

*The optimal control for* (14) *is unique and is given by*

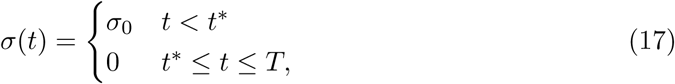

where

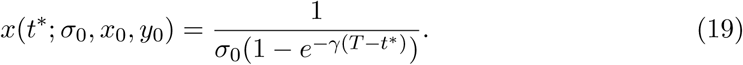

*and otherwise t*^*^ *is the unique solution of*

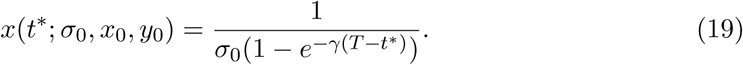

### Proof.

First, suppose *x*(0) ≤ 1/*σ*_0_. The claimed optimal control gives *x*(*T*) = *x*_0_, whereas any other control will give *x*(*T*) < *x*_0_. Similarly, we see from (2) that the optimal control gives *y*(*T*) = *e*^−γT^*y*_0_ and any other control will lead to a larger value of *y*(*T*). Since *x*_∞_ is a decreasing function of *y* and (for *x* < 1/*σ*_0_) an increasing function of *x*, the proposed control is optimal in this case.

Now suppose *x*(0) > 1/*σ*_0_. We reformulate the objective as follows. From (7) we see that *x*_∞_ is a strictly monotone increasing function of *μ*, so that maximizing *x*_∞_ is equivalent to maximizing *μ*. Now

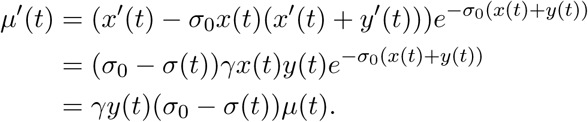

Thus

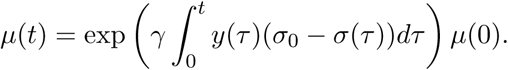

Thus, maximizing *x*_∞_(*T*) is equivalent to maximizing

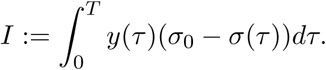

From Theorem 2 we have that

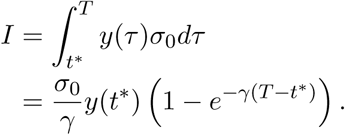

Differentiating with respect to *t*^*^ gives

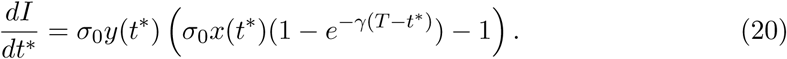

If this condition in (18) is satisfied then this has no zero and 1 is maximized by taking *t*^*^ = 0. If the condition in (18) is not satisfied, then setting the right hand side of (20) equal to zero yields the condition (19). By checking the second derivative, it is easily confirmed that this is indeed a maximum. □

We remark that the above result apparently cannot be obtained via standard sufficiency conditions based on Pontryagin’s maximum principle, due to the nonconvexity of the right hand side of the SIR system (2).

Some optimal solutions for particular instances of (14) are shown in Figures 4 and 5, all with the same initial data and parameters *β*, *γ* but with different final times *T*. Allowing for a longer intervention (larger *T*) makes it possible to reach a more optimal value of *x*_∞_.

**Figure 4:**
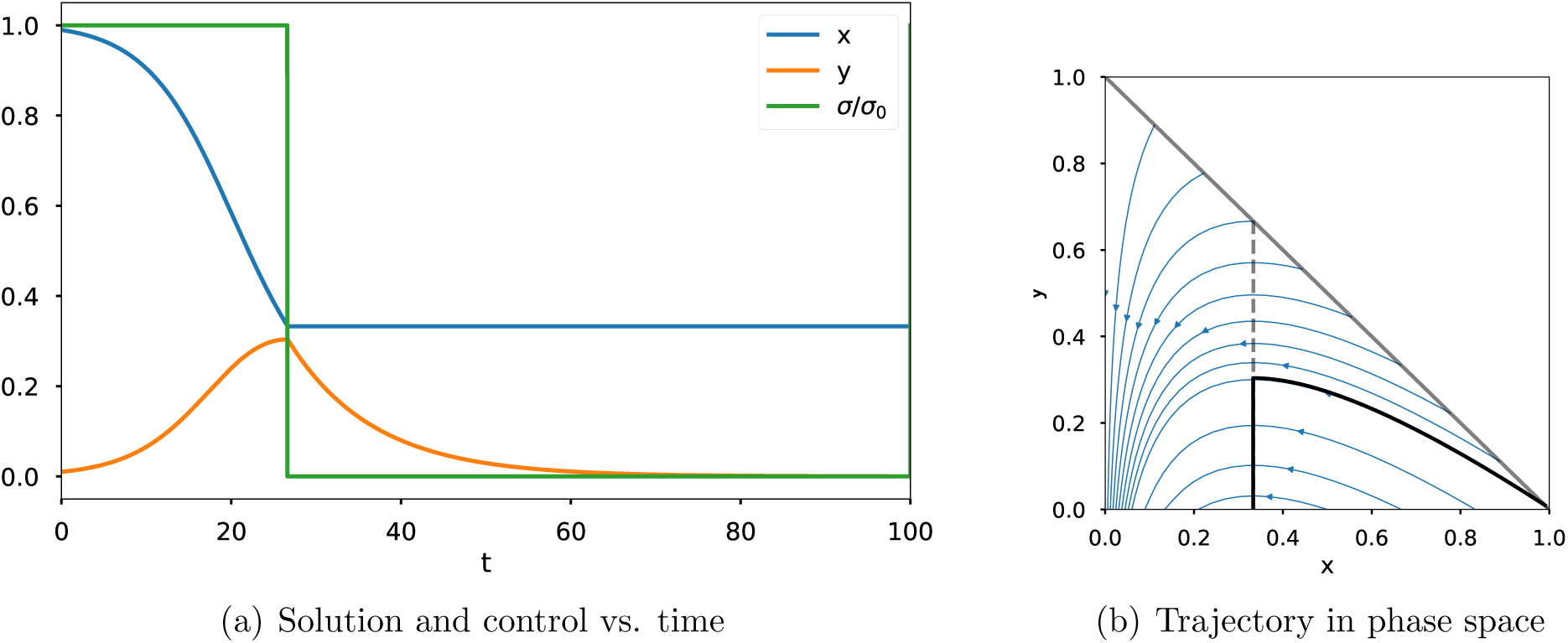
Typical optimal solution. Here (*x*(0)*,y*(0)) = (0.99, 0.01), *β* = 0.3, and *γ* = 0.1.

**Figure 5:**
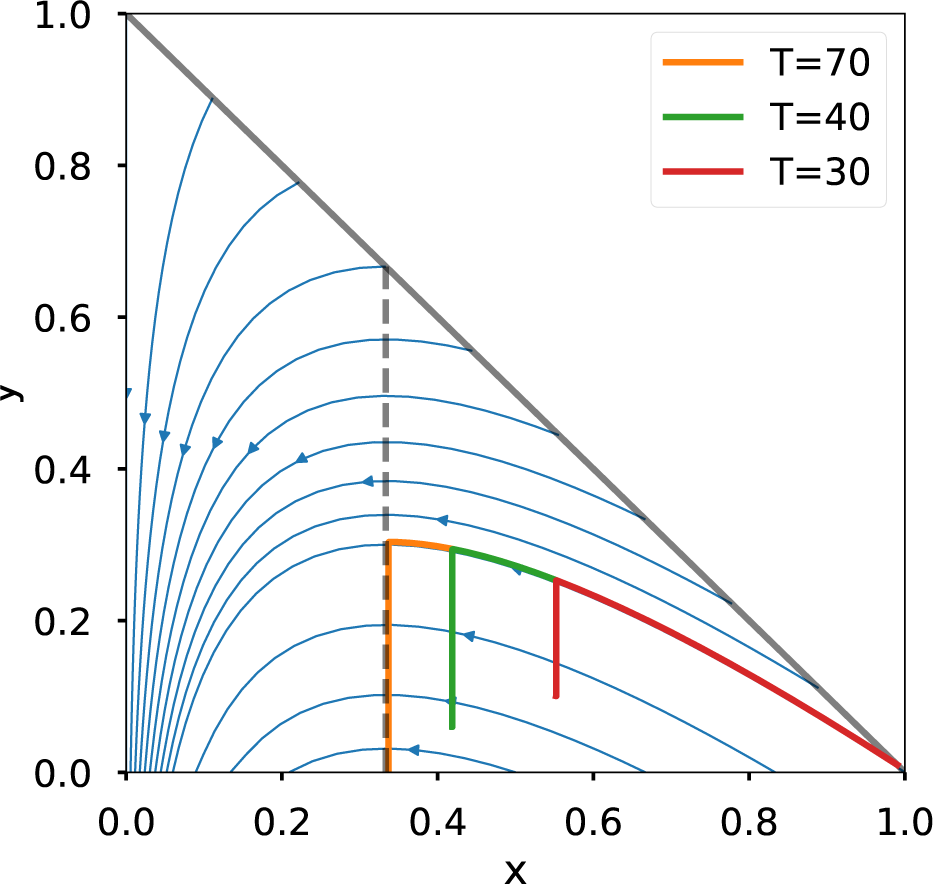
Optimal solutions starting from the same point (0.99, 0.01) but with different final times. A larger value of *T* allows the system to reach a more optimal state. For all solutions, *β* = 0.3 and *γ* = 0.1.

In real-world scenarios, it may not be possible to apply the maximum control *σ*(*t*) = 0. Suppose that in place of (3) we impose *σ*_min_ ≤ *σ*(*t*) ≤ *σ*_0_. In this case the optimal control is still bang-bang with a single switching time. In Figure 6, we show an optimal solution when *σ*(*t*) ≥ 0.4*σ*_0_ is imposed.

**Figure 6:**
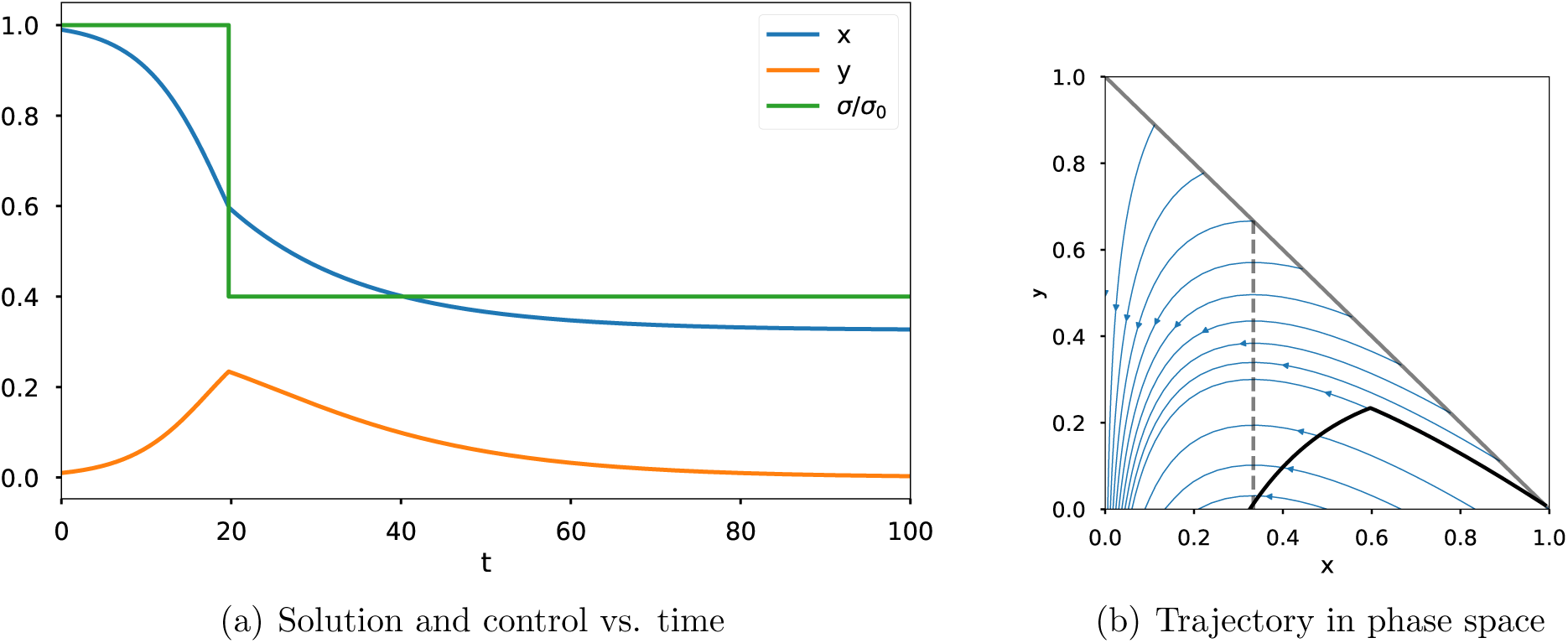
Optimal solutions with *σ*(*t*) ≥ 0.4*σ*_0_. Here (*x*(0), *y*(0)) = (0.99,0.01), *β* = 0.3, *γ* = 0.1, and *T* = 100.

The result above can also be obtained via the Hamilton-Jacobi-Bellman (HJB) equation for (14). Here we sketch this approach. The HJB equation for u(*x*, *y*, *t*) can be written

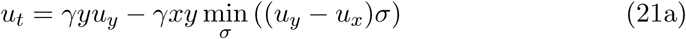

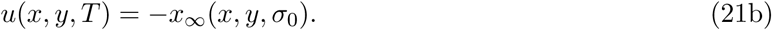

The required minimum is obtained by taking

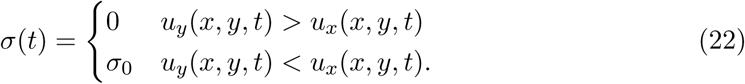

From (7) we see that *u_y_*(*x*, *y*, *T*) > *u_x_*(*x*, *y*, *T*) for all (*x*, *y*). Thus for small enough values of *T* — *t*, the solution of (21) satisfies

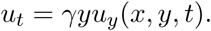

The solution of this hyperbolic PDE is

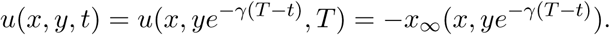

Thus, for small enough *T* − *t*,

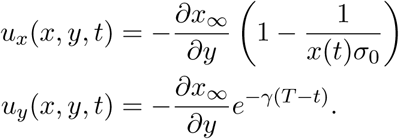

According to (22), the optimal control value will switch when *u_x_* = *u_y_*, which leads to (19). Meanwhile, substituting (22) in (21) in the case *u_y_* < *u_x_* yields the linear hyperbolic PDE

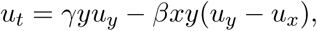

whose characteristics are just the trajectories of the SIR system (1) illustrated in Figure 1, which are also contours of *x*_∞_. It can be shown that once *u_y_* − *u_x_* < 0, this inequality will continue to hold along each such characteristic.

## 4 Optimal control with *L* ≠ 0

We now consider the case of a non-zero Lagrangian, which allows us to account for factors like the economic cost of intervention or heightened risks caused by hospital overflow. We formulate the Hamiltion-Jacobi-Bellman (HJB) equation for this problem and apply an upwind numerical method to compute approximate solutions. The numerical solutions obtained via the HJB equation have also been checked in each case against solutions of the BVP given in Section 2.3, and found to agree within numerical errors.

### 4.1 Quadratic running cost of control

We now attempt to account for the economic cost of intervention. Quantification of the cost of measures like closing schools and businesses is a challenging problem in economic modeling, and well outside the scope of the present work. Based on the general idea that both the cost and the marginal cost will increase with the degree of contact reduction, we take for simplicity

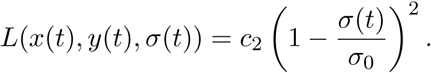

The HJB equation for (6) is then

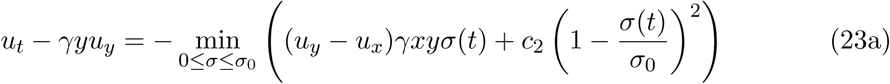

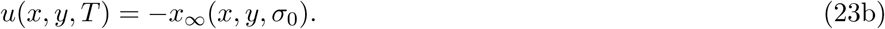

The minimum in (23a) is obtained with

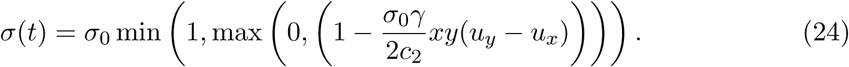

We approximate the solution of (23)-(24) using the first-order upwind finite difference method:

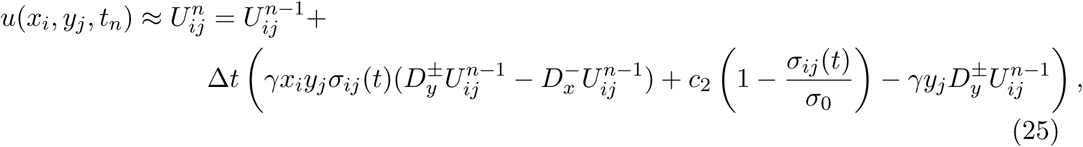

where

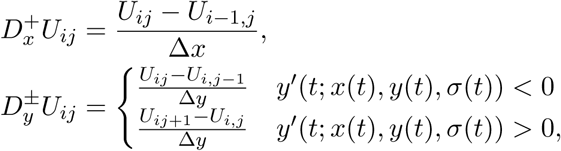

and *σ_ij_*(*t*) is given by a centered difference discretization of (24). Note that the upwind *x*-direction is always to the left, since *x*′(*t*) ≤ 0.

Numerical solutions for a range of values of *c*_2_ are shown in Figure 7. The values of *c*_2_ used here are chosen merely to illustrate the range of possible behaviors. Notice that the strength of the control *σ*(*t*) at early times varies non-monotonically with *c*_2_, first increasing and then decreasing as *c*_2_ is reduced. Indeed, the optimal control *σ*(*t*) over the initial time interval is simply *σ*_0_ in both limits *c*_2_ → ∞ and *c*_2_ → 0.

**Figure 7:**
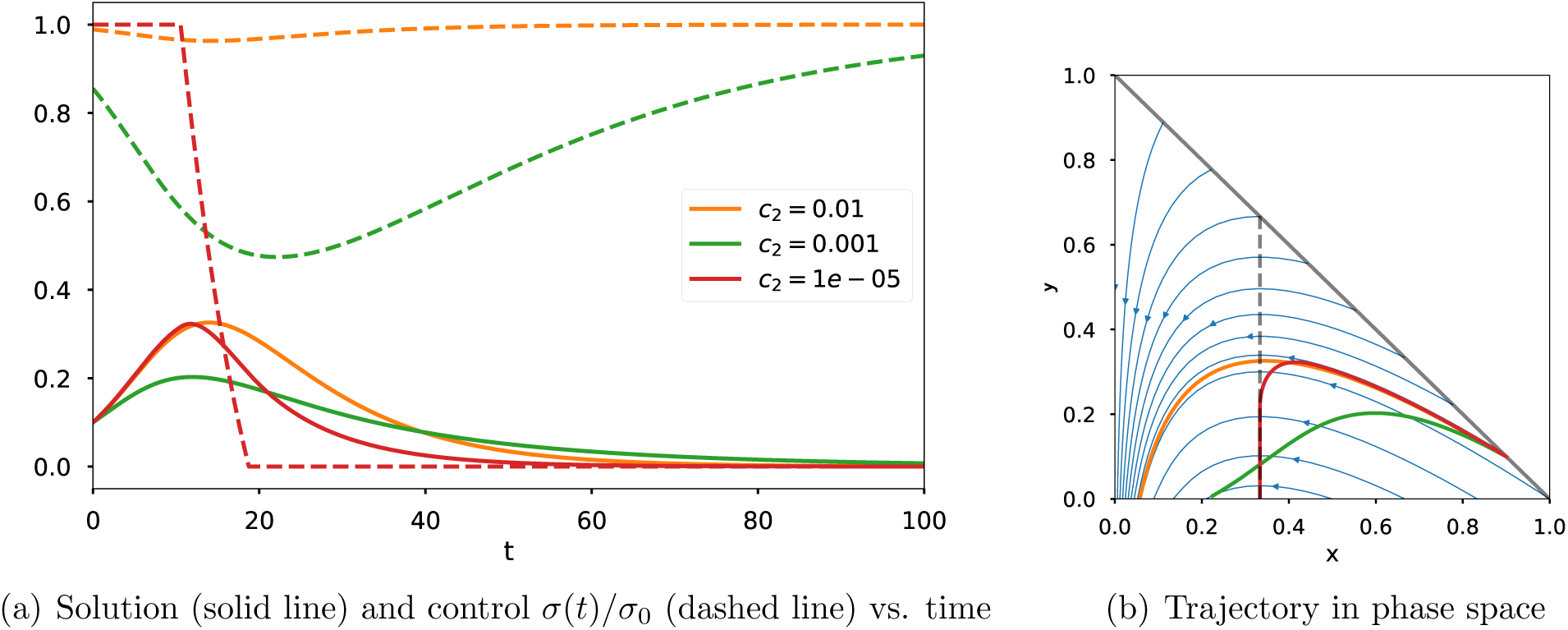
Optimal solutions with different running cost. Here (*x*(0), *y*(0)) = (0.9, 0.1), *β* = 0.3, *γ* = 0.1, and *T* = 100.

### 4.2 Minimizing hospital overflow

The optimal solutions above may be unsatisfactory in practice, since the number of people simultaneously infected at certain times may be too great for all of them to receive adequate medical care. This is a major concern with respect to the current COVID-19 crisis. A natural objective is to keep the number of infected below some threshold, corresponding for instance to the number of hospital beds. We thus consider the Lagrangian

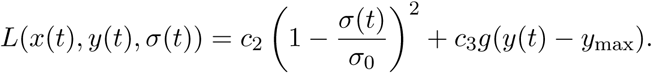

Here *y*_max_ is the maximum number of hospital beds. The HJB equation is then

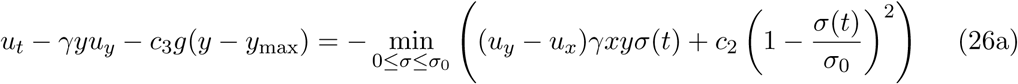

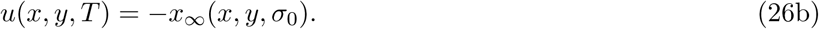

The control that achieves the minimum in (26a) is again given by (24). The function *g*(*υ*) should be nearly zero for *υ* < 0 and increase in an approximately linear fashion for *υ* > 0. For the purpose of having a tractable control problem, it is also desirable that *g* be differentiable. We take

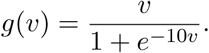

Figures 8 and 9 show examples of solutions. Again, we choose parameter values that demonstrate the range of qualitative behaviors. In both examples, the cost of control is scaled by *c*_2_ = 10^−3^. In Figure 8, a higher cost for hospital overflow is applied, with *c*_3_ = 10. As might be expected, *y*(*t*) is generally kept below *y*_max_ (which is set to 0.1). The control is initially off, then turns on to avoid hospital overflow, and then turns off again. While the control is applied, it is maintained at a level that keeps the value of *y*(*t*) nearly constant in time.

**Figure 8:**
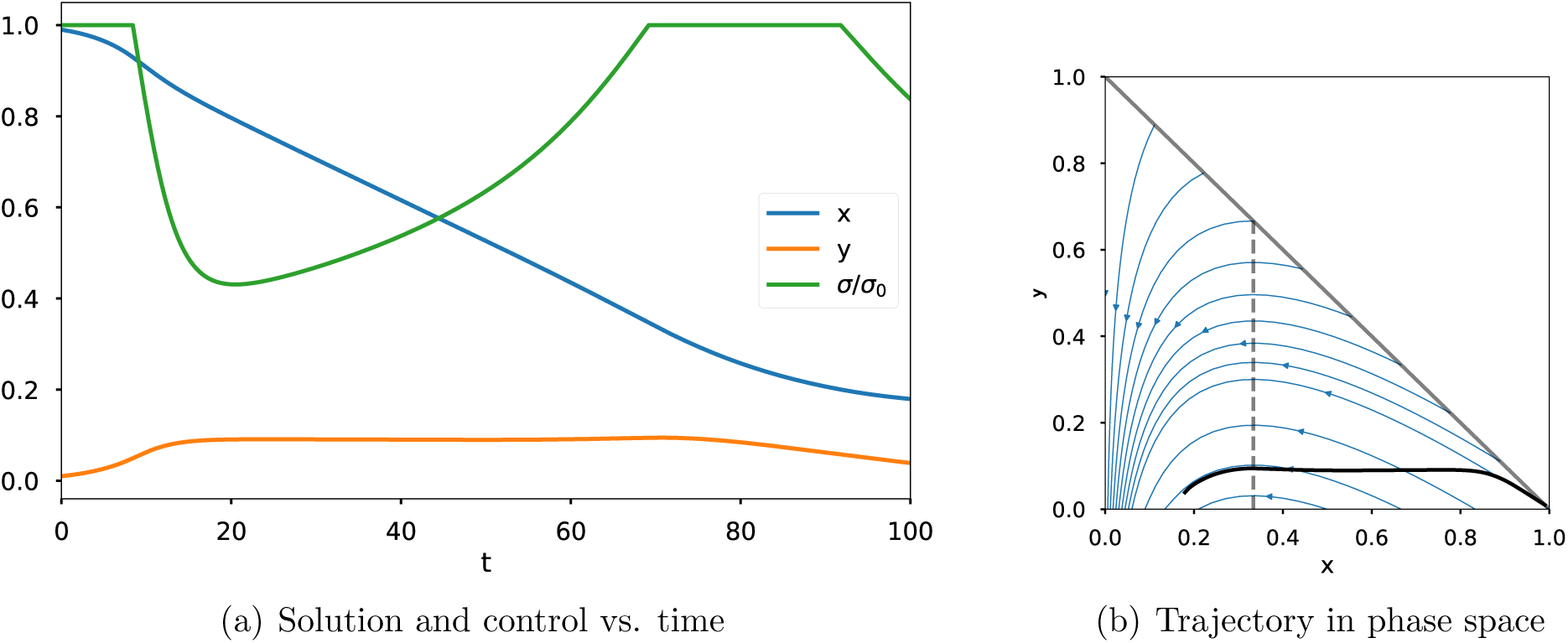
Optimal solutions with cost for hospital overflow. Here (*x*(0),*y*(0)) = (0.99, 0.01), *β* = 0.3, *γ* = 0.1, *T* = 100, and *y*_max_ = 0.1. In the cost function, we take c_2_ = 10^−3^ and *c*_3_ = 10.

**Figure 9:**
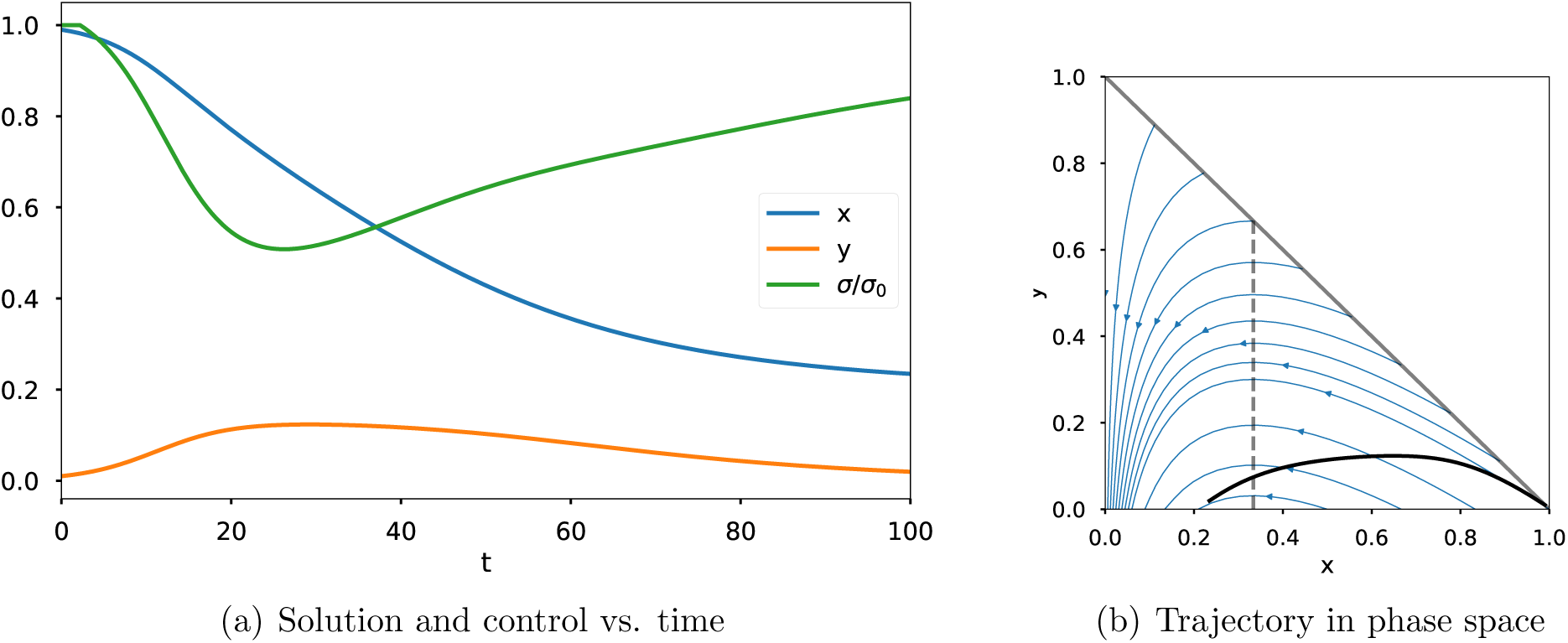
Optimal solutions with smaller cost for hospital overflow. Here (*x*(0),*y*(0)) = (0.99,0.01), *β* = 0.3, *γ* = 0.1, *T* = 100, and *y*_max_ = 0.1. In the cost function, we take *c*_2_ = 10^−3^ and *c*_3_ = 1.

Figure 9 shows another example scenario in which the cost of hospital overflow is smaller, with *c*_3_ = 1. In this case the hospital capacity is significantly exceeded for a short time, and the control is kept on until the final time, but the epidemiological overshoot is significantly reduced compared to the previous solution.

## 5 Application to the COVID-19 pandemic

The main goal of this work has been a mathematical investigation of optimal controls for the SIR model with a controlled rate of contact, as presented in the previous sections. We now present a brief illustration of the results in practical terms through application to the current COVID-19 pandemic. This application is imprecise, for several reasons: the SIR model is one of the simplest epidemiological models, and assumes homogeneous mixing among a population; the current state of susceptible and infected persons is not accurately known; and the parameters of the disease itself (i.e. *γ*,σ_0_) are still quite uncertain. The examples in this section should be viewed only as illustrations of a few possible scenarios, and not an exhaustive or detailed study.

We take the infectious period γ^−1^ = 10 days, and the basic reproduction number *σ*_0_ = 2.5, based on recent estimates [23, 16]. To make the results easy to interpret, we use a fixed terminal cost of *c*_1_*z*_∞_, where we have introduced an additional scaling constant. Taking *c*_1_ = *αN*, where *N* is the total population being modeled and *α* is the infection fatality ratio, then this cost is the expected number of lives lost. Since *z*_∞_ = 1 − *x*_∞_, this is merely a rescaling of the terminal cost used throughout this work. We take *α* ≈ 0.006 based on recent estimates [23, 19, 24].

We seek reasonable order-of-magnitude estimates for *c*_2_ and *c*_3_. The value of *c*_3_/*N* should be equal to the increase in probability of a given infected person dying because of the lack of medical care. We take *c*_3_ = *Nη*, where the fatality ratio in the absence of medical care is *α* + *η*. In the absence of any relevant data, we take *η* ≈ 2*α*, giving *c*_3_ = 0.012*N*. For *y*_max_ we take values from the United States, where there are about 3 hospital beds per 1000 people, and two-thirds of them are typically occupied. Since it is estimated that about 5% of COVID-19 cases are hospitalized [23], this gives *y*_max_ = 0.02*N*.

Any attempt to quantify the cost of an intervention in human lives is bound to be contentious. Whether we consider the value of a human life to be in intrinsic personal value or extrinsic economic value, we can view the cost of intervention as a reduction of the value of human lives during the intervention period. We take *c*_2_ = *N ϵ /d* where *d* ≈ 10^4^ is the number of days in a human life (more precisely, the average number of days remaining in a life claimed by the disease) and 1 − *ϵ* is the relative value of a day spent in full isolation (*σ* = 0) compared to a day without intervention. Taking e = 0.2, we have *c*_2_ = 2 × 10^−5^*N*.

Since all terms in the cost function are proportional to *N*, we take *N* = 1 without loss of generality. Results for the parameter values given above are shown in Figure 10. We see that the optimal control corresponds to a level of intervention that becomes more strict as the epidemic grows, and is gradually relaxed as the epidemic subsides. Most importantly, and in agreement with results from the examples in earlier sections, the strongest control is applied around the time of peak infection and shortly thereafter. The maximum hospital capacity is significantly exceeded, with the maximum value of y around 0.08. It seems plausible that this level of infection may still be manageable with local surges of care facilities and staff, like those that have already been carried out in practice for COVID-19. There is also a noticeable (but greatly reduced relative to the uncontrolled case) epidemiological overshoot.

**Figure 10:**
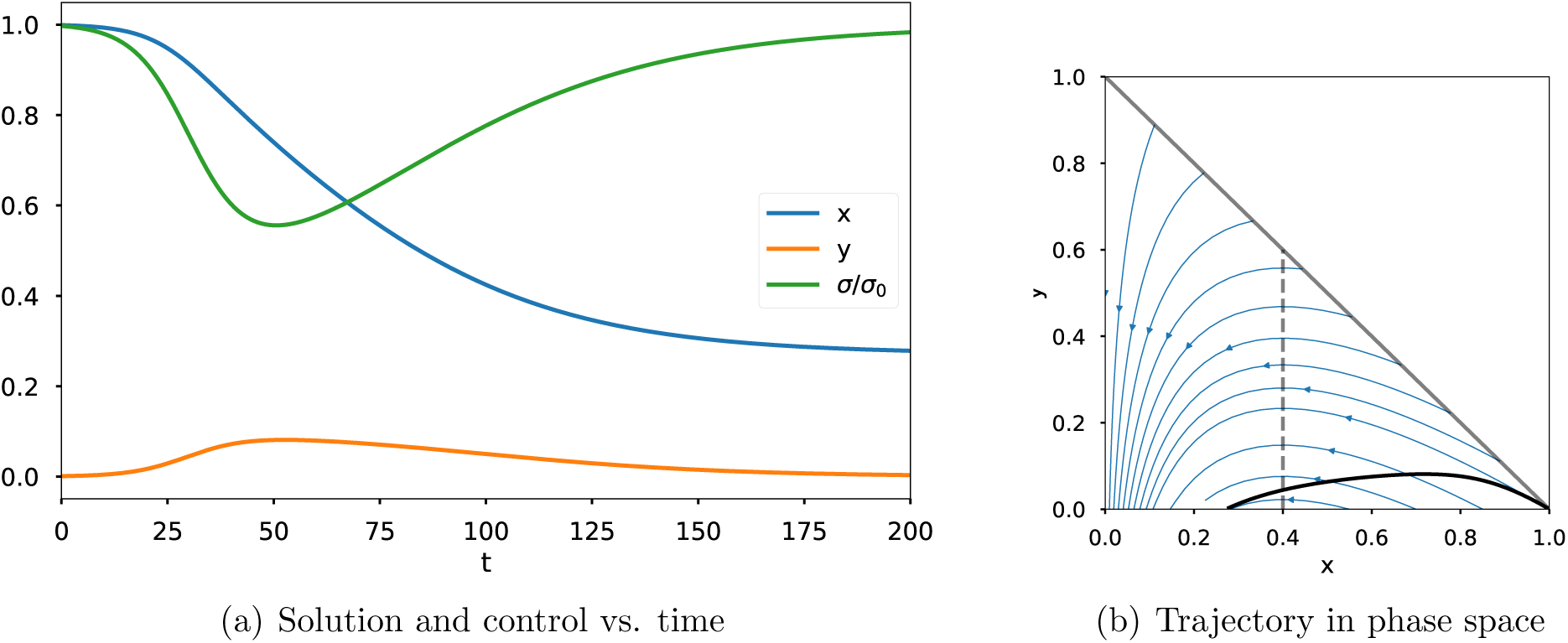
Optimal control for COVID-19 with *σ*_0_ = 2.5, *γ* = 0.1, *α* = 0.006, *η* = 0.012, *ϵ* = 0.2, *d* = 10^4^, *T* = 200, and (*x*(0), *y*(0)) = (0.999, 0.001).

An alternative scenario is shown in Figure 11, in which we have assumed a fatality ratio and a value of *η* that are twice as large (in line with the highest estimates of the infection fatality ratio), as well as taking a smaller cost of intervention with *ϵ* = 0.05. These parameters lead to stronger intervention, especially in the later phases of the epidemic. The result is almost no epidemiological overshoot and a reduction in maximum simultaneous infected (to around 0.06).

**Figure 11:**
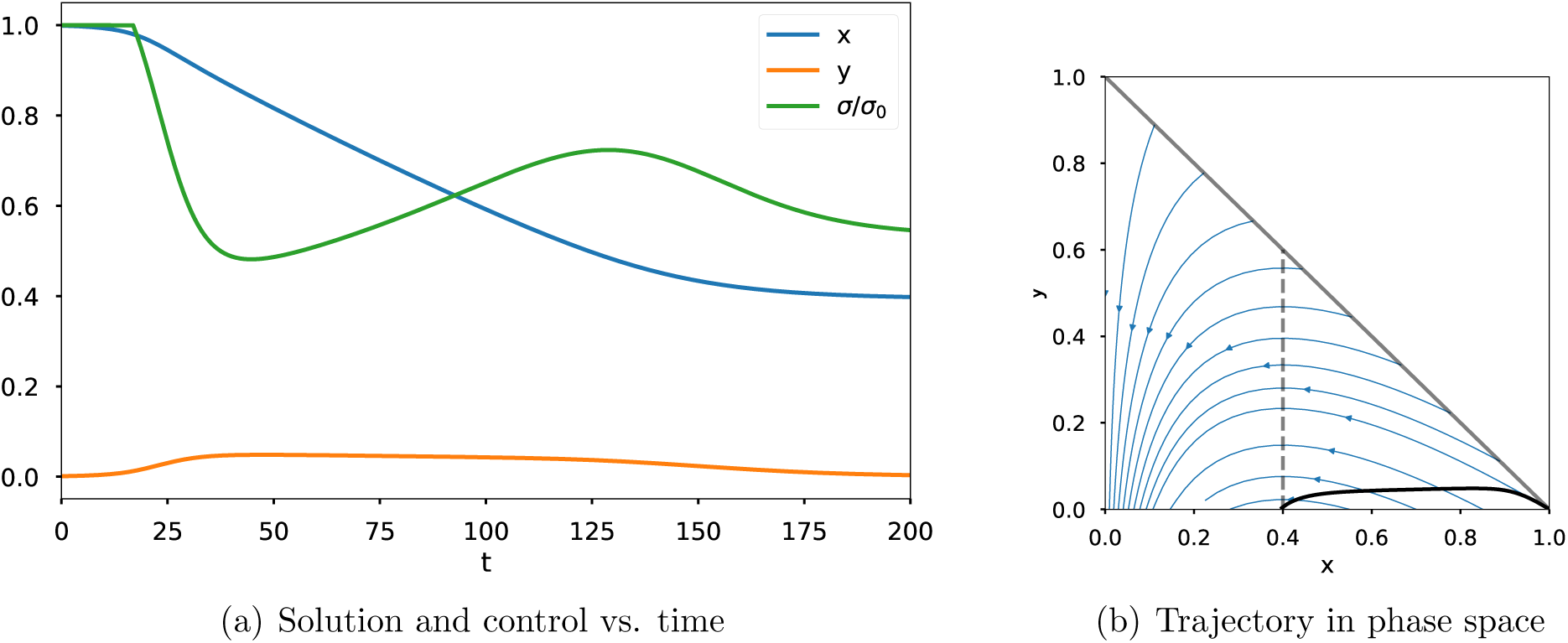
Optimal control for COVID-19 with *σ*_0_ = 2.5, *γ* = 0.1, *α* = 0.012, *η* = 0.024, *ϵ* = 0.05, *d* = 10^4^, *T* = 200, and (*x*(0), *y*(0)) = (0.999, 0.001).

Finally, in Figure 12, we repeat the first scenario but increase the cost of control by taking *ϵ* = 0.5. In this case a more mild control is applied, peaking at about 35% contact reduction and concentrated around the time of the infection peak. Both the epidemiological overshoot and the maximum infected are worse, compared to the previous two scenarios.

**Figure 12:**
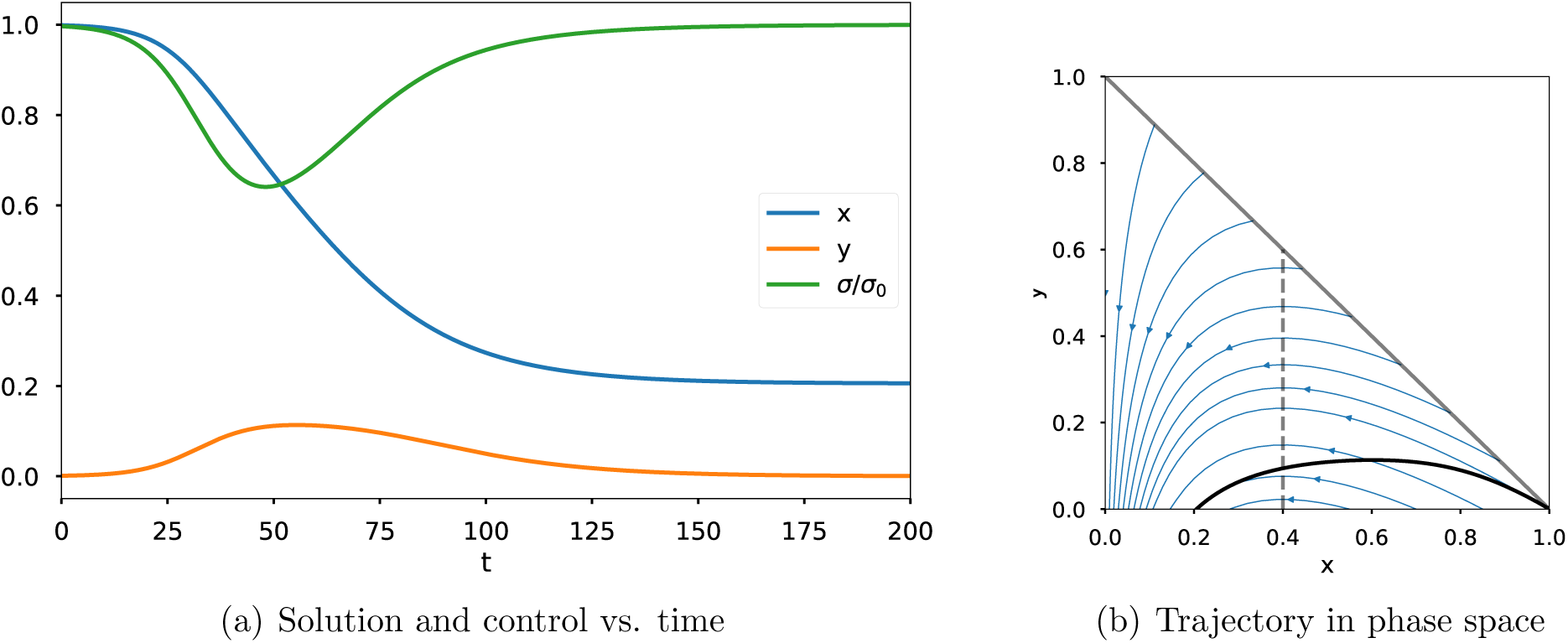
Optimal control for COVID-19 with *σ*_0_ = 2.5, *γ* = 0.1, *α* = *η* = 0.006, *ϵ* = 0.5, *d* = 10^4^, *T* = 2000, and (*x*(0), *y*(0)) = (0.999, 0.001).

## 6 Conclusion

We have studied, for an SIR model with a control on the rate of contact, the problem of minimizing the eventually infected population in the long-time limit, when the control can be applied only up to a finite time. In the absence of any cost of intervention, the optimal strategy is to apply no control until a certain switching time, and then apply maximum control. We have also considered other objective functions that include a running cost of control and a penalty for large numbers of simultaneous infections.

Contrary to simple intuition, it is not optimal to impose the maximum level of intervention from the earliest possible time. But real-world studies have supported this observation; a too-strong intervention may simply lead to a strong second wave of infection after the intervention is lifted, and not significantly reduce epidemiological overshoot [3]. On the other hand, intervention that starts too late or is lifted too soon may also have a negligible effect on total mortality [3, 9, 17]. The idea that intervention should possibly be delayed in order to increase its effect was also found in [2], although the objective and optimal policy found there differ from the present work.

Real-world application of the strategies derived here would require precise knowledge of the disease parameters, the current state of the population, and the quantitative effect of specific NPIs, none of which are readily available (or indeed capable of being characterized by a single number). Nevertheless, the general results obtained here may provide insight into what optimal intervention strategies and their consequences may look like. A major shortcoming of the present work is the assumption of a uniform mortality rate across all individuals in a population. We are currently working on a generalization of this work that includes different levels of risk for different subpopulations.

## Data Availability

No data is associated with this article. Some related code is available at https://github.com/ketch/SIR_control

## References

[1] F. B. Agusto. Optimal isolation control strategies and cost-effectiveness analysis of a two-strain avian influenza model. Biosystems, 113(3):155–164, 2013.

[2] PG Ballard, NG Bean, and JV Ross. Intervention to maximise the probability of epidemic fade-out. Mathematical Biosciences, 293:1–10, 2017.

[3] Martin CJ Bootsma and Neil M Ferguson. The effect of public health measures on the 1918 influenza pandemic in US cities. Proceedings of the National Academy of Sciences, 104(18):7588–7593, 2007.

[4] Neil M. Ferguson, Derek A. T. Cummings, Simon Cauchemez, Christophe Fraser, Steven Riley, Aronrag Meeyai, Sopon Iamsirithaworn, and Donald S Burke. Strategies for containing an emerging influenza pandemic in Southeast Asia. Nature, 437(7056):209–214, 2005.

[5] K Renee Fister, Suzanne Lenhart, and Joseph Scott McNally. Optimizing chemotherapy in an HIV model. Electronic Journal of Differential Equations, 1998(32):1–12, 1998.

[6] Wendell H Fleming and Raymond W Rishel. Deterministic and stochastic optimal control, volume 1. Springer Science & Business Media, 2012.

[7] David Greenhalgh. Some results on optimal control applied to epidemics. Mathematical Biosciences, 88(2):125–158, 1988.

[8] Tiberiu Harko, Francisco S. N. Lobo, and M. K. Mak. Exact analytical solutions of the susceptible-infected-recovered (SIR) epidemic model and of the SIR model with equal death and birth rates. Applied Mathematics and Computation, 236:184–194, 2014.

[9] Richard J. Hatchett, Carter E. Mecher, and Marc Lipsitch. Public health interventions and epidemic intensity during the 1918 influenza pandemic. Proceedings of the National Academy of Sciences, 104(18):7582–7587, 2007.

[10] Herbert W Hethcote. The mathematics of infectious diseases. SIAM review, 42(4):599–653, 2000.

[11] E Jung, Suzanne Lenhart, and Z Feng. Optimal control of treatments in a two-strain tuberculosis model. Discrete & Continuous Dynamical Systems-B, 2(4):473, 2002.

[12] T. K. Kar and Ashim Batabyal. Stability analysis and optimal control of an SIR epidemic model with vaccination. Biosystems, 104(2–3):127–135, 2011.

[13] William Ogilvy Kermack and Anderson G. McKendrick. A contribution to the mathematical theory of epidemics. Proceedings of the Royal Society of London. Series A, Containing papers of a mathematical and physical character, 115(772):700–721, 1927.

[14] Denise Kirschner, Suzanne Lenhart, and Steve Serbin. Optimal control of the chemotherapy of HIV. Journal of Mathematical Biology, 35(7):775–792, 1997.

[15] Suzanne Lenhart and John T Workman. Optimal control applied to biological models. CRC press, 2007.

[16] Ying Liu, Albert A Gayle, Annelies Wilder-Smith, and Joacim Rocklöv. The reproductive number of COVID-19 is higher compared to SARS coronavirus. Journal of Travel Medicine, 2020.

[17] Howard Markel, Harvey B Lipman, J Alexander Navarro, Alexandra Sloan, Joseph R Michalsen, Alexandra Minna Stern, and Martin S Cetron. Nonpharmaceutical interventions implemented by US cities during the 1918–1919 influenza pandemic. JAMA, 298(6):644–654, 2007.

[18] Anthony G. Pakes. Lambert’s W meets Kermack-McKendrick epidemics. IMA Journal of Applied Mathematics, 80(5):1368–1386, 2015.

[19] Timothy W Russell, Joel Hellewell, Christopher I Jarvis, Kevin Van-Zandvoort, Sam Abbott, Ruwan Ratnayake, Stefan Flasche, Rosalind M Eggo, Adam J Kucharski, CMMID nCov working group, et al. Estimating the infection and case fatality ratio for COVID-19 using age-adjusted data from the outbreak on the Diamond Princess cruise ship. *medRxiv*, 2020.

[20] Mohammad A. Safi and Abba B. Gumel. Dynamics of a model with quarantine-adjusted incidence and quarantine of susceptible individuals. Journal of Mathematical Analysis and Applications, 399(2):565–575, 2013.

[21] Oluwaseun Sharomi and Tufail Malik. Optimal control in epidemiology. Annals of Operations Research, 251(1–2):55–71, 2017.

[22] Haoxuan Sun, Yumou Qiu, Han Yan, Yaxuan Huang, Yuru Zhu, and Song Xi Chen. Tracking and predicting COVID-19 epidemic in China Mainland. *medRxiv*, 2020.

[23] Robert Verity, Lucy C Okell, Ilaria Dorigatti, Peter Winskill, Charles Whittaker, Natsuko Imai, Gina Cuomo-Dannenburg, Hayley Thompson, Patrick GT Walker, Han Fu, et al. Estimates of the severity of coronavirus disease 2019: a model-based analysis. The Lancet Infectious Diseases, 2020.

[24] Joseph T Wu, Kathy Leung, Mary Bushman, Nishant Kishore, Rene Niehus, Pablo M de Salazar, Benjamin J Cowling, Marc Lipsitch, and Gabriel M Leung. Estimating clinical severity of COVID-19 from the transmission dynamics in Wuhan, China. Nature Medicine, pages 1–5, 2020.

[25] Xiefei Yan and Yun Zou. Optimal and sub-optimal quarantine and isolation control in SARS epidemics. Mathematical and Computer Modelling, 47(1–2):235–245, 2008.

